# Automated Segmentation of Post-Surgical Resection Cavities on MRI in Focal Epilepsy: a MELD Study

**DOI:** 10.64898/2026.02.26.26347093

**Authors:** Jieun Seo, Mathilde Ripart, Helene Kaas, Ben Sinclair, Lucy Vivash, Merran R. Courtney, Terence J O’Brien, Siby Gopinath, Harilal Parasuram, Sedat Kandemirli, Natally Alarab, Lillian Lai, Marcus Likeman, Kai Zhang, Jiajie Mo, Georgian Ciobotaru, James Galea, Philip Sequeiros-Peggs, Khalid Hamandi, Hua Xie, Venkata Sita Priyanka Illapani, William D. Gaillard, Nathan T. Cohen, Alexander G. Weil, Florence Henrichon-Goulet, Kenza S. Lahlou, Aristides Hadjinicolaou, Agustín Ibáñez, Gonzalo M. Rojas-Costa, Horst Urbach, Lara Bücheler, Marcel Heers, Adrián Valls Carbó, Rafael Toledano, Giulia Nobile, Costanza Parodi, Domenico Tortora, Alessandro Consales, Antonella Riva, Mariasavina Severino, Martin Tisdall, Felice D’Arco, Kshitij Mankad, Aswin Chari, Maria H. Eriksson, Rory J. Piper, J. Helen Cross, Torsten Baldeweg, Sofia González-Ortiz, Jose Pariente, Nuria Bargalló, Yawu Liu, Reetta Kälviäinen, Carmen Barba, Matteo Lenge, Renzo Guerrini, Masaki Iwasaki, Daichi Sone, Hiroyuki Maki, Tomoki Imokawa, Noriko Sato, Julien Jung, Francisco Sepulveda, Daniel Mansilla, Andres Goycoolea, Ingeborg Lopez, Antonio Napolitano, Alessandro De Benedictis, Luca De Palma, Maria Camilla Rossi-Espagnet, Nikolaos Kondylidis, Kostakis Gkiatis, Kyriakos Garganis, Joshua Pepper, Stefano Seri, John S. Duncan, Clarissa L. Yasuda, Lucas Scárdua-Silva, Marina K. M. Alvim, Fernando Cendes, Antonio G. Gennari, Ruth O’Gorman Tuura, Georgia Ramantani, Mariam Josyula, Joel Stein, Nishant Sinha, Kate Davis, R. Edward Hogan, Luigi Maccotta, Sophie Adler, Konrad Wagstyl

## Abstract

**Objective:** Quantitative assessment of extent of tissue resection following epilepsy surgery requires accurate delineation of the resection cavity on postoperative MRI. Current methods for resection cavity masking are time-consuming and labour-intensive, while existing automated approaches exhibit variable segmentation accuracy, particularly on extra-temporal resections. We developed MELD-PostOp, a deep learning tool trained and evaluated on a large, heterogeneous cohort to automatically segment resection cavities.

**Methods:** The study included 1.5 and 3T postoperative 3D T1-weighted MRI images from the Multicentre Epilepsy Lesion Detection (MELD) project (n_subjects_=969, 27 centres) and from the EPISURG dataset (n=133). The cohort included children and adults, alongside a range of resection locations, pathologies, and MRI characteristics. Resection cavities were individually segmented in 285 subjects and used to train an nnU-Net prototype model. The prototype model was used to generate an additional 680 resection masks, which were subsequently quality-controlled, edited and combined with the original 285 to train the final MELD-PostOp model (n=965). A Stratified (STC; n=50) and Independent Test Cohort (ITC; n=87) were withheld for model evaluation. Performance was evaluated using Dice Similarity Coefficient (DSC), 95th percentile Hausdorff distance (HD95), number of predicted clusters and inference runtime; and compared against established tools (Epic-CHOP and ResectVol).

**Results:** MELD-PostOp achieved a median DSC of 0.85 and HD95 of 3.61 on the combined test cohort, outperforming Epic-CHOP (DSC 0.68, HD95 9.54) and ResectVol (DSC 0.66, HD95 12.07), with significant improvements seen in both temporal and especially extra-temporal resections. The model detected 99% (135/137) of resection cavities. MELD-PostOp runtime was 17s per MRI, compared to 612s (ResectVol) and 3205s (Epic-CHOP). MELD-PostOp performance remained high across clinical and imaging subgroups (median DSC > 0.8).

**Significance:** MELD-PostOp provides an accurate, efficient and generalisable solution for postoperative resection cavity segmentation using only postoperative MRI scans. This open-source tool facilitates large-scale quantitative analysis to define what tissue is essential to resect for optimal epilepsy surgical outcomes.

**Key points:**

- MELD-PostOp automatically segments resection cavities from a postoperative MRI scan in 17 seconds, a 100-200x faster runtime than existing tools
- The model was trained on 965 annotated T1-weighted postoperative scans from sites worldwide, supporting its generalisation across diverse clinical and imaging settings.
- The model achieves high segmentation accuracy (DSC > 0.8), with consistently high-quality segmentations across age groups, sex, image resolutions, and resection locations.

## 1. Introduction

Epilepsy surgery is an established and effective treatment for patients with drug-resistant focal epilepsy^1^ caused by structural aetiologies, for which computational techniques may help to optimise resection strategies. Complete resection of the epileptogenic zone is necessary for achieving postoperative seizure freedom.^2,3^ However, the resection extent must be balanced with limiting the removal of functional tissue to minimise neurocognitive consequences.^4^ For MRI visible lesions, postoperative MRI plays a key role in assessing resection extent and analysing its association with post-surgical seizure freedom and neurocognitive outcomes. In patients undergoing a reoperation due to seizure recurrence, approximately 60% of cases were attributed to incomplete resection of the epileptogenic zone, often resulting from lesion mislocalisation, residual epileptogenic tissue, or surgical limitations imposed by proximity to eloquent cortex.^5^ However, the evaluation of resection extent frequently relies on qualitative rather than quantitative assessments, i.e. radiologists visually assess whether the lesion has been completely resected.^5^ Such visual assessments are subject to interrater variability, dependent on reader expertise, and are usually binary (complete vs. incomplete), limiting the ability to assess whether the proportion of the structural lesion resected or resection of specific regions of the epileptogenic zone are important in determining post-operative outcomes. Accurate delineation of the resection cavity is essential for research into understanding and improving surgical outcomes.

Quantitative masks of resection extent can be generated from postoperative MRI scans with manual, semi-automated and fully-automated approaches.^6–9^ Manual masking by experts remains the gold-standard, but is time-consuming and varies between raters.^10^ Semi-automated methods accelerate this process by using computational algorithms to propagate user-generated segmentation prompts into 3D masks. These are then manually reviewed and updated.^6,7^ Whilst faster, even semi-automated segmentation of images limits the capacity to scale analysis to large cohorts with hundreds or thousands of subjects and is still subject to inter-rater variability.

Several studies have proposed automated approaches for resection cavity segmentation following epilepsy surgery.^8,9,11,12^ These approaches mainly fall into two categories. The first involves coregistration of preoperative and postoperative scans, followed by voxel-wise subtraction or intensity difference mapping.^8,9^ Epic-CHOP and ResectVol are two open source examples of this approach which are both built on the Statistical Parametric Mapping (SPM) toolbox. The second category encompasses deep learning (DL) methods, typically based on convolutional neural network (CNN) architectures,^13^ trained to segment the resection cavity using real or synthetic examples of postoperative scans with corresponding ground-truth labels. RESSEG is one example of a DL approach trained using the open EPISURG dataset. EPISURG is an adult cohort of primarily temporal-lobe resections.^11,14,15^

A number of limitations hamper the utility of both SPM-based and DL methods. SPM-based methods require accurate coregistration and tissue segmentation of pre and postoperative images. This multistage process is time-consuming and introduces potential sources of error. DL methods on the other hand are dependent on large sample sizes which must accurately capture the true heterogeneity of the target population in order to generalise to independent external validation cohorts. For resection cavity masking, DL methods have, to date, only been trained on data from one or two centres. In empirical validation, existing SPM methods outperformed DL approaches. However, both methods failed to detect the resection on 12-16% and 54-56% of cases respectively. Notably, all methods performed less well on extra-temporal lesions, failing on 15-30% (SPM-based) and 85% (DL-based) of cases.^16^ These limitations highlight the need for a robust, generalisable segmentation framework capable of accurately delineating resection cavities using only postoperative MRI data.

To address this, we collected a large, heterogeneous multicentre dataset of postoperative three-dimensional (3D) T1-weighted (T1w; 1.5T or 3T) MRI images from paediatric and adult patients with a range of epilepsy pathologies distributed across the brain. We introduce a scalable annotation strategy, combining manual labelling and active learning to define the resection cavities. We trained an nnU-Net, a self-configuring model that selects the optimal architecture and hyperparameters and has demonstrated state-of-the-art performance across a range of biomedical segmentation tasks.^17^ We evaluated our model, MELD-PostOp, on two withheld test cohorts and compared performance to existing approaches, Epic-CHOP^9^ and ResectVol^8^. MELD-PostOp, a robust and generalisable model for segmentation of resection cavities on post-operative T1w scans is provided open-source for independent validation and research into optimising surgical outcomes.

## 2. Materials and methods

### 2.1 Study population

#### 2.1.1 Datasets and inclusion criteria

This study used anonymised MRI data collected as part of the Multi-centre Epilepsy Lesion Detection (MELD) Focal Epilepsies project (https://meldproject.github.io/), which has Health Research Authority approval (IRAS: 301863), and the open-source EPISURG dataset. The MELD Focal Epilepsies dataset (v1.0, January 2025) includes 1090 participants with post-operative MRI scans from 27 epilepsy centres. Each participating centre obtained local ethical approval to retrieve, anonymise and share retrospective, routinely acquired clinical imaging data and clinical variables with the central MELD research team. Patients were included if they underwent lesionectomy or lobectomy for focal epilepsy and had a postoperative 3D T1w brain MRI scan. Excluded patients included those who had undergone disconnection procedures, laser ablation or thermocoagulation.

Clinical variables included age, sex, pathology, and surgical procedure. Age at surgery, or age at preoperative scan, dependent on data availability, was classified as paediatric (<18 years), or adult (≥18 years). Pathologies were defined by post-operative histopathology or MRI diagnosis, dependent on data availability. These were grouped as 1) malformation of cortical development (MCD), including focal cortical dysplasia (FCD), periventricular nodular heterotopia, polymicrogyria and mild malformation of cortical development with oligodendroglial hyperplasia and epilepsy (MOGHE), 2) cavernoma, 3) hippocampal sclerosis (HS), 4) low-grade epilepsy associated tumour (LEAT), 5) dual pathology, 6) other, or 7) unknown. Dual pathology referred to cases in which more than one distinct pathological lesion was identified. “Other” pathologies included cortical gliosis, hypothalamic hamartoma (HH), or normal MRI findings without a specific histopathological diagnosis. Unknown indicated cases where pathological information was unavailable or not specified in the anonymised clinical data received.

In addition, the open-source EPISURG dataset (n=430),^14,15^ a clinical dataset of postoperative T1w MRI scans of adult epilepsy patients who underwent resective surgery, was incorporated. Available clinical variables included the surgical procedure and operated hemisphere. EPISURG patients were included if they had a 3D T1w postoperative scan with corresponding resection cavity masks.

#### 2.1.2 Cohort creation

A total of 1102 patients met the inclusion criteria: 969 patients from the MELD Focal Epilepsies dataset and 133 patients from the EPISURG dataset. These patients were divided into 4 cohorts: 1) Prototype Cohort (n=285), 2) MELD-PostOp Train Cohort (n=965), 3) Stratified Test Cohort (STC; n=50) and 4) Independent Test Cohort (ITC; n=87) (Figure 2C).

##### 2.1.2.1 The Prototype Cohort

To train the model, a fully quality-controlled set of resection cavity masks for all 965 patients in the MELD-PostOp Train Cohort need to be created. To accelerate the manual process, a subset Prototype Cohort was semi-automatically segmented in a two-stage process (n=285). First, nnInteractive was used via the napari graphical user interface. This semi-automated tool uses a pretrained network to refine users’ prompts to generate an initial delineation of resection cavities.^7,18^ The masks were visually quality controlled and manually refined when necessary using ITK-SNAP.^6^ Manual resection cavity masks for all 285 scans were generated by members of the team (n=152, JS, RP, BH, EN) or downloaded as part of the EPISURG dataset (n=133). Second, all resection cavity masks underwent subsequent visual quality control (QC) and, where necessary, correction by a single rater (JS) prior to incorporation into the Prototype Cohort (Figure 1). These labelled images were used to train the Prototype nnU-Net model (see section 2.2 for nnU-Net).

**Figure 1.**
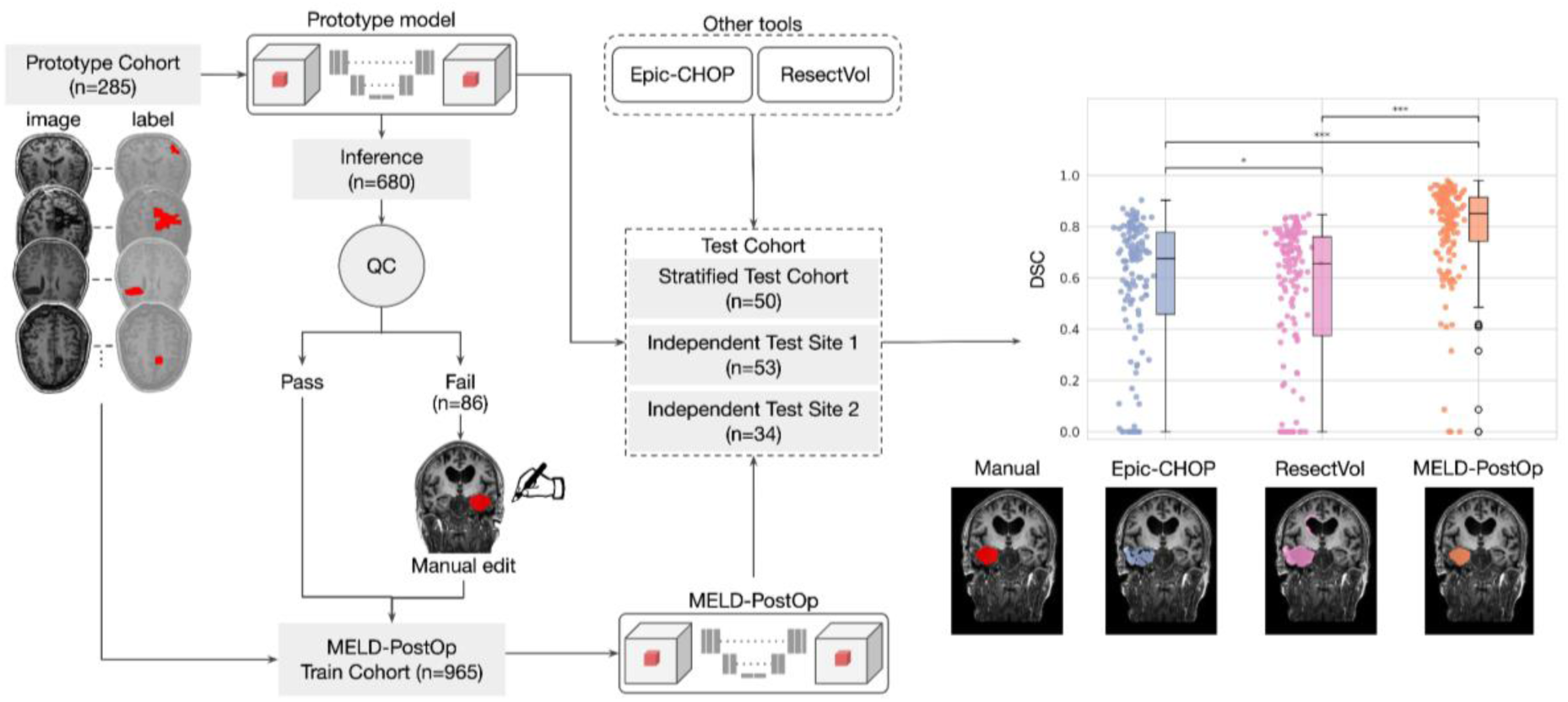
MELD-PostOp overview. The Prototype Cohort (n=285) was used to train the prototype model. The prototype model was applied to an inference cohort (n=680). All predictions underwent visual quality control (QC). Failed cases were manually refined using nnInteractive and ITK-SNAP. The masks that passed QC and manually edited masks were combined with the Prototype Cohort to form the MELD-PostOp Train Cohort (n=965), which was used to train the final MELD-PostOp model. Model performance was evaluated on the Test Cohorts. MELD-PostOp performance was compared against Epic-CHOP and ResectVol.

##### 2.1.2.2. The MELD-PostOp Train Cohort

The MELD-PostOp Train Cohort combines subjects in the Prototype Train Cohort (n=285) with the remaining MELD Focal Epilepsies patients (n=680). To generate labels for these 680 scans, the Prototype model was used to automatically predict resection cavities. Multiview QC images were reviewed for all 680 680 predicted cavities (Supp. Fig 1, 2).

Cases that failed QC (n=86) were manually refined using ITK-SNAP. The approved and manually-edited segmentations were combined with the Prototype Train Cohort to form the MELD-PostOp Train Cohort (n=965). This MELD-PostOp Train Cohort was used to train the final MELD-PostOp model.

##### 2.1.2.3 The Stratified Test Cohort (STC)

To test differences in performance across clinical and demographic factors, 50 subjects from the full MELD Focal Epilepsies cohort were assigned to a STC that was withheld from all training and optimisation experiments. Subjects were randomly selected, stratifying by site, age and pathology.

##### 2.1.2.4 The Independent Test Cohort (ITC)

To evaluate MELD-PostOp performance on data from an unseen scanner, two MELD Focal Epilepsies sites were withheld as the ITC (total n=87). Sites were chosen as they were large (n=53; n=34) with a range of pathologies and ages.

Ground-truth segmentation masks for all subjects both test cohorts were created with the same semi-automated segmentation workflow used for the Prototype Cohort.

### 2.2 nnU-Net resection cavity models

The Prototype and MELD-PostOp models were trained using the nnU-Net framework with the 3D full-resolution configuration, with postoperative T1w 3D MRI scan as input, and resection cavity masks as the target segmentation. Five-fold cross-validation was performed, with 80% of the data allocated for training and 20% for validation in each model. Folds were randomly stratified to maintain balanced label representation across the training and validation sets. The final model was an ensemble averaging predictions from the five models. All hyperparameters were automatically selected by nnU-Net, except the number of epochs which was increased to 2000.

### 2.3 Other automated segmentation tools

Two automated epilepsy surgery resection tools, Epic-CHOP^9^ and ResectVol,^8^ were selected as leading existing methods.^16^ Both tools run within Statistical Parametric Mapping (SPM12).^19^ Briefly they automatically predict the resection cavity by coregistering the preoperative and post-operative scans and computing voxel-wise differences, with methods detailed in the original papers^8, 9^.

Source code for each tool was downloaded from:

- Epic-CHOP: https://github.com/iBrain-Lab/EPIC-CHOP
- ResectVol: https://www.lniunicamp.com/resectvol

### 2.4 Evaluation of automated segmentation tools on Test Cohorts

MELD-PostOp, Epic-CHOP and ResectVol were evaluated on the STC and ITC. Resection cavity masks were automatically segmented by each model. Evaluation metrics included: dice similarity coefficient (DSC) and 95th percentile Hausdorff distance (HD95) between the ground truth mask and each automatically segmented mask, the number of predicted clusters, and model runtime.

DSC captures overall agreement between ground truth and predicted masks and was computed as: 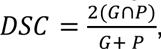 where G is the binary ground truth mask, and P is the binary automatically segmented masks. One limitation of DSC is that it tends to be lower for smaller objects. HD95 captures the distance between the boundaries of the objects and was computed as: *HD*95 = *max*(*percentile*_95_(*D*(*P* → *G*)), *percentile*_95_(*D*(*G* → *P*))), where D is the shortest distance from the surface of one mask to the surface of the other. Model performance was summarised using the median DSC and HD95 with associated interquartile ranges (IQR). The Wilcoxon signed-rank test was applied to compare the median DSC between models. Also calculated were the proportions of cases with DSC > 0, i.e. sensitivity of the model to detect a resection cavity, and DSC > 0.5, which represents the proportion of cases with a meaningful overlap^16^.

Clusters were defined as contiguous connected voxels, categorised as True positive (TP) if there was overlap with the ground truth mask and false positives (FP) if not. The median number of FP clusters per image with IQR was reported. Positive predictive value (PPV) was calculated as: 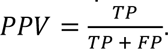

Mean runtime per image was measured by recording the runtime in seconds for each model applied to the same set of 10 postoperative images and calculating the mean and standard deviation.

### 2.5 Sub-group analysis of models’ performances

Performance in the test cohorts (n=137) was stratified by age group, sex, histopathology, resection lobe, surgical side, postoperative MRI field strength, and isotropic image resolution. Due to sample size limitations, for subgroups with more than two categories, the category with the highest median DSC was compared against all remaining categories (e.g. Temporal vs Extra-temporal) using Mann-Whitney U tests.

Group-stratified performance of MELD-PostOp was also quantified combining the predictions on validation cohorts (i.e. cross-validation predictions) with predictions on the test cohorts. The EPISURG dataset was excluded from this analysis since patient demographics and clinical variables were unavailable.

Median and IQR for DSC and HD95 were computed within each subgroup. Mann-Whitney U tests were used for comparisons between binary variables (e.g. sex, adult vs paediatric). Kruskal-Wallis tests were used to test for between-group heterogeneity for stratifiers with three or more categories (i.e., pathology and lobe), followed by pairwise Mann-Whitney U tests. The relationship between DSC/HD95 and resection cavity volume was analysed using Spearman’s rank correlation test. Bonferroni adjustment was used to correct p-values for multiple subgroup comparisons (n=8), and were considered statistically significant if *p_adjusted_* < 0.05.

To group scan acquisitions, MRI scans were considered isotropic if the resolution in all three dimensions was within +/-0.1, else anisotropic.

Postoperative images and their corresponding ground truth masks were non-linearly registered to MNI-ICBM152 space^21^ (Figure 2d). Overlap with lobes from the Cerebrum Atlas (CerebrA)^22^ and the resection cavity masks was computed. The lobar label with the highest DSC was used to define the resection lobe and hemisphere. Cavities not overlapping a cortical lobe (e.g. hypothalamic hamartoma) were grouped into the “other” category. SynthSeg-derived segmentations were used to quality control quality control co-registrations. Initial registrations were computed using ANTsPy. For those with mean DSC below 0.5, registrations were recomputed with EasyReg^24,25^.

**Figure 2.**
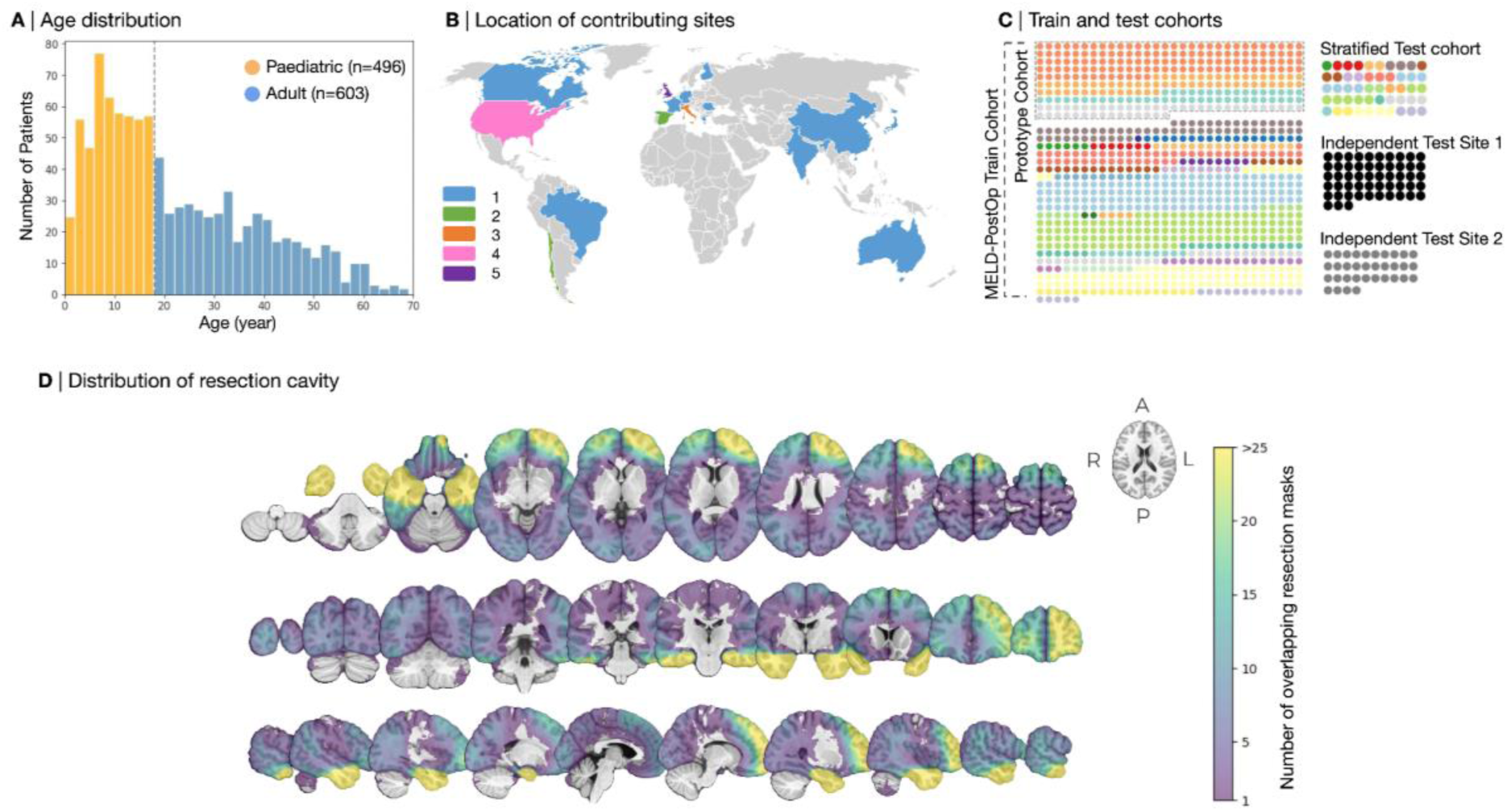
Visualisation of patient characteristics in the study population. (A) Age distribution and classification of patients into paediatric (<18 years) and adult (≥18 years) groups. (B) Geographical distribution of contributing sites, with each colour indicating the number of participating sites from a country. A total of 28 sites (MELD sites and EPISURG) across 17 countries contributed. (C) Cohort composition. Prototype Cohort (n=285) included patients from three MELD sites and the EPISURG dataset. MELD-PostOp Train Cohort (n=965) incorporated Prototype Cohort and additional patients from 25 MELD sites. Randomly selected patients from 25 MELD sites formed the Stratified Test Cohort (n=50), and two MELD sites served as the Independent Test Cohort (n=53; n=34). (D) Volumetric density map of manually segmented resection cavity masks in MNI space, demonstrating widespread coverage with the highest overlap density, i.e. the most frequently operated locations, in the temporal and left frontal lobes.

### 2.6 Code and model availability

All code for model design, development and statistical analyses alongside the MELD-PostOp model is available at: MELDProject Github.

## 3. Results

### 3.1 Study population

#### 3.1.1 Patient characteristics

969 patients from 27 MELD sites and 133 patients from the EPISURG dataset were included in the study (total n=1102). Demographic and clinical characteristics of the cohort are illustrated in Figure 2 and summarised in Table 1. The entire cohort comprised 45.0% paediatric and 54.7% adult patients, with 45.7% males and 42.2% females. Histopathological diagnoses were heterogeneous, with MCD being the most prevalent pathological group (37.8%), followed by LEAT (17.6%) and HS (17.2%). The temporal lobe was the most frequently resected (55.6%), followed by the frontal lobe (25.3%), (Figure 2D). Resections were evenly distributed between left (51.5%) and right (48.5%) hemispheres. The majority of postoperative MRI scans were acquired at 3T (76.7%) and were of isotropic resolution (72.6%). The majority of scans were obtained within the first postoperative year (median 0 years [IQR 0-1], range 0-15 years).

**Table 1.** Clinical and imaging characteristics of the study population (n=1,102). MCD, malformation of cortical development; FCD, focal cortical dysplasia; HS, hippocampal sclerosis; LEAT, low-grade epilepsy-associated tumour; MRI, magnetic resonance imaging

|  | Number of patients (n) | Percentage (%) |
| --- | --- | --- |
| Age |  |  |
| Paediatric | 496 | 45.0 |
| Adult | 603 | 54.7 |
| Unknown | 3 | 0.3 |
| Sex |  |  |
| Male | 504 | 45.7 |
| Female | 465 | 45.2 |
| Unknown | 133 | 12.1 |
| Pathology |  |  |
| MCD | 421 | 38.2 |
| HS | 190 | 17.2 |
| LEAT | 194 | 17.6 |
| Cavernoma | 48 | 4.4 |
| Dual pathology | 83 | 7.5 |
| Other | 23 | 2.1 |
| Unknown | 143 | 13.0 |
| Location of resection |  |  |
| Temporal | 613 | 55.6 |
| Frontal | 279 | 25.3 |
| Occipital | 39 | 3.5 |
| Parietal | 105 | 9.5 |
| Limbic | 32 | 2.9 |
| Insular | 9 | 0.8 |
| Other | 25 | 2.3 |
| Surgical side |  |  |
| Left | 567 | 51.5 |
| Right | 535 | 48.5 |
| MRI field strength |  |  |
| 3T | 845 | 76.7 |
| 1.5T | 124 | 11.3 |
| Unknown | 133 | 12.1 |
| Isotropy |  |  |
| Isotropic | 800 | 72.6 |
| Anisotropic | 302 | 27.4 |

### 3.2 Overall model performances

Model performances on the STC and ITC across 3 models (MELD-PostOp, Epic-CHOP and ResectVol) are summarised in Table 2 and illustrated in Figure 3. MELD-PostOp achieved a median DSC of 0.85 across the two cohorts, significantly higher than Epic-CHOP and ResectVol (p < 0.001), and a meaningful overlap (DSC > 0.5) in 93.4% of patients.

**Figure 3.**
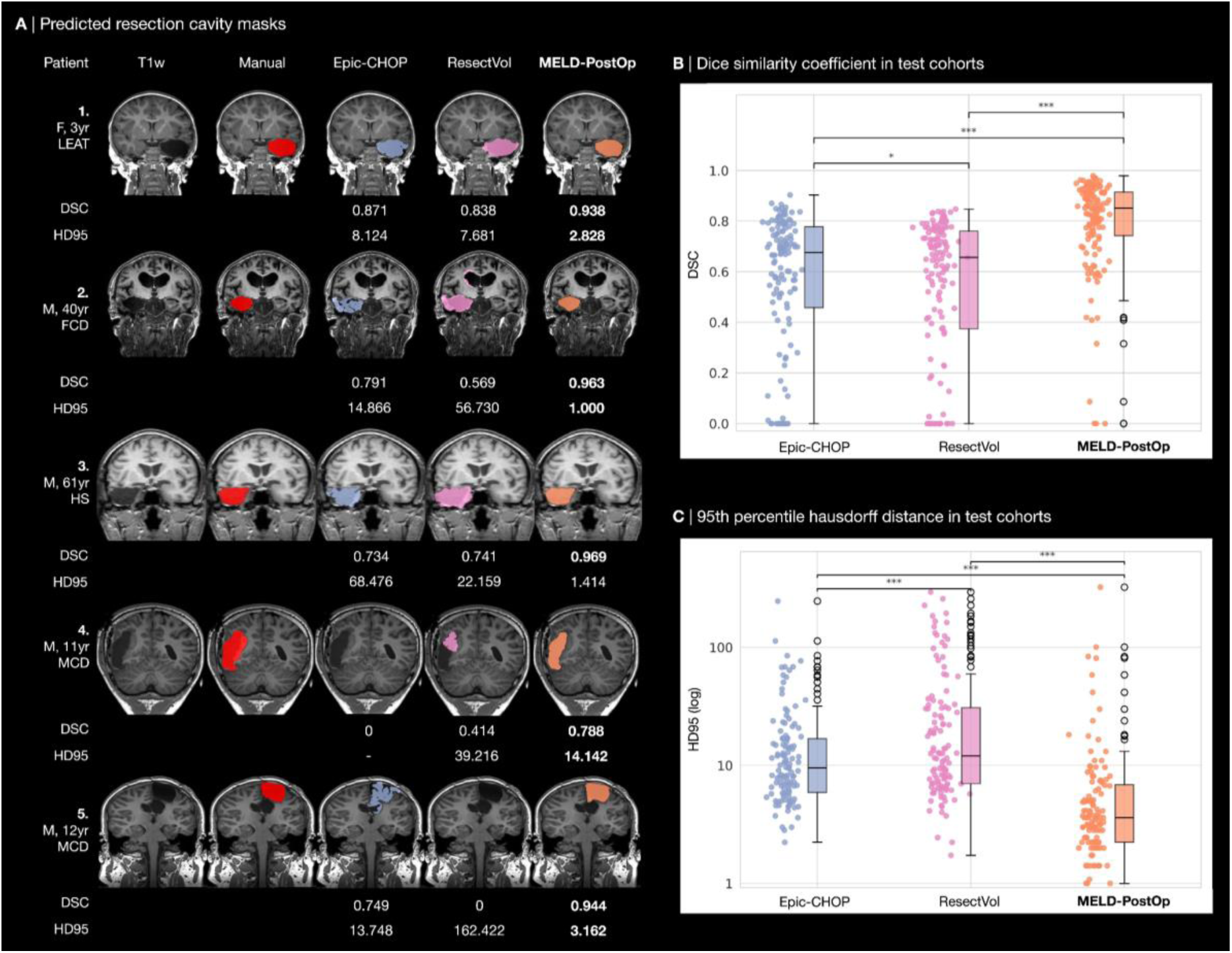
[A] Raw postoperative T1w MRI scans, ground truth manual resection masks, and automated segmentation from Epic-CHOP, ResectVol and MELD-PostOp models. The number displayed beneath each predicted mask indicates the Dice Similarity Coefficient (DSC) and the 95th percentile Hausdorff distance (HD95) relative to the manual mask. [B, C] Box and scatter plots of DSCs (B) and HD95 (C) across models in the combined test cohorts (n=137). MELD-PostOp achieved a significantly higher median DSC and lower HD95 than Epic-CHOP and ResectVol (*p < 0.001)*. LEAT, low grade epilepsy associated tumour; FCD, focal cortical dysplasia; HS, hippocampal sclerosis; MCD, malformations of cortical development; HD95, 95th percentile Hausdorff distance

**Table 2.** Model performances of automated resection cavity segmentation models on the Stratified Test Cohort and Independent Test Cohort. Performance metrics are presented as median dice similarity coefficient (DSC), DSC > 0, DSC > 0.5, median HD95, PPV, number of FP clusters, and mean runtime per image. IQR, interquartile range; HD95, 95th percentile Hausdorff distance; PPV, positive predictive value; SD, standard deviation

| | | Median DSC<br>[IQR] | DSC > 0 (%) | DSC > 0.5 (%) | Median HD95<br>[IQR] | PPV | Median FP<br>clusters [IQR] | Mean runtime ±<br>SD (sec/image) |
| --- | --- | --- | --- | --- | --- | --- | --- | --- |
| Epic-CHOP | Stratified | 0.61 [0.33-0.76] | 41/50 (82.0) | 34/50 (68.0) | 10.84 [5.85-22.47] | 0.09 | 3 [1-12] | 3205 ± 876 |
|  | Independent | 0.70 [0.51-0.79] | 80/87 (92.0) | 65/87 (74.7) | 9.54 [6.66-15.22] | 0.25 | 3 [1-6] |  |
|  | Combined | 0.68 [0.48-0.78] | 121/137 (88.3) | 99/137 (72.3) | 9.54 [5.89-16.85] | 0.17 | 11 [5-29] |  |
| ResectVol | Stratified | 0.62 [0.14-0.74] | 39/50 (78.0) | 32/50 (64.0) | 10.39 [6.48-32.93] | 0.35 | 1 [0-3] | 612 ± 160 |
|  | Independent | 0.68 [0.42-0.76] | 77/87 (88.5) | 61/87 (70.1) | 12.69 [7.89-31.93] | 0.20 | 1 [0-5] |  |
|  | Combined | 0.66 [0.37-0.76] | 116/137 (84.7) | 93/137 (67.9) | 12.07 [7.00-30.86] | 0.24 | 1 [0-4] |  |
| MELD-PostOp | Stratified | 0.83 [0.68-0.89] | 50/50 (100) | 46/50 (92.0) | 3.87 [3.00-7.14] | 0.95 | 0 [0-0] | 17 ± 5 |
|  | Independent | 0.87 [0.76-0.94] | 85/87 (97.7) | 82/87 (97.3) | 3.32 [2.24-6.78] | 0.93 | 0 [0-0] |  |
|  | Combined | 0.85 [0.74-0.92] | 135/137 (98.5) | 128/137 (93.4) | 3.61 [2.24-6.87] | 0.93 | 0 [0-0] |  |

MELD-PostOp successfully identified all resection cavities (DSC > 0) in the STC (100% sensitivity) and 85 out of 87 resection cavities in the ITC (97.7% sensitivity). The two cases MELD-PostOp failed to segment were in the smallest 6.6% of resections (1.03 and 2.28 cm^3^). In comparison, Epic-CHOP failed to segment a resection cavity in 16, and ResectVol in 21 cases.

MELD-PostOp had a significantly higher positive predictive value (PPV=0.93, p < 0.001) compared to Epic-CHOP (PPV=0.17) and ResectVol (PPV=0.24), with false positive predictions in only 6% across both test cohorts. Additionally, MELD-PostOp had a median HD95 of 3.61, significantly lower than Epic-CHOP (9.54) and ResectVol (12.07) (p < 0.001).

Runtime analysis showed MELD-PostOp had a mean processing time of 17 seconds per image (SD, 5s), compared to 612 seconds (i.e. 10 minutes) (SD, 160s) for ResectVol and 3205 seconds (i.e. 53 minutes) (SD, 876s) for Epic-CHOP (*p* < 0.001).

Model outputs are visualised for a number of example cases (Figure 3A). Three of these patients underwent temporal lobe resec tions for different temporal aetiologies (LEAT, FCD, and HS). All models successfully segmented the temporal lobe resection cavitie s. However, MELD-PostOp achieved higher DSC values exceeding 0.94 compared to less than 0.88 for Epic-CHOP and ResectVol. The fourth and fifth examples represent paediatric patients with non-temporal resections due to underlying MCD. MELD-PostOp accurately segmented these resection cavities, reaching DSCs of 0.79 and 0.94, respectively. In contrast, Epic-CHOP failed to produce a segmentation for example 4, while ResectVol failed on example 5.

### 3.3 Subgroup analysis of models’ performances

Comparing models’ performances in subgroups (Table 3), MELD-PostOp achieved significantly higher segmentation performance in both temporal and extra-temporal resections compared with Epic-CHOP and ResectVol. Across all three models, DSC were significantly higher in temporal than in extra-temporal cases. MELD-PostOp exhibited the smallest reduction in performance between temporal and extra-temporal resections (median DSC difference=0.07), compared to Epic-CHOP and ResectVol (median DSC difference 0.13 and 0.18, respectively). MELD-PostOp did not significantly differ in performance across other subgroups.

**Table 3.** Models’ performances breakdown across clinical and imaging variables (n=137). Median DSCdice [IQR] and median HD95 [IQR] for Epic-CHOP, ResectVol and MELD-PostOp models categorised by patient age, sex, pathology, resection location, surgical laterality, postoperative MRI field strength, and isotropy. *, p <0.05; **, p< 0.01; ***, p<0.001; ns, not significant; HD95, 95th percentile Hausdorff distance; HS, hippocampal sclerosis; MRI, magnetic resonance imaging

|  | Epic-CHOP |  | ResectVol |  | MELD-PostOp |  |
| --- | --- | --- | --- | --- | --- | --- |
|  | DSC | HD95 | DSC | HD95 | DSC | HD95 |
| Age |  |  |  |  |  |  |
| Paediatric | 0.68 [0.41-0.79] | 13.9 [8.2-21.8] | 0.61 [0.00-0.74] | 23.0 [7.6-56.6] | 0.84 [0.78-0.91] | 3.6 [2.2-7.6] |
| Adults | 0.68 [0.52-0.75] | 8.3 [5.8-14.9] | 0.68 [0.50-0.76] | 11.5 [7.0-30.2] | 0.86 [0.74-0.92] | 3.6 [2.3-6.8] |
| Sex |  |  |  |  |  |  |
| Male | 0.67 [0.48-0.76] | 11.1 [5.8-18.3] | 0.62 [0.35-0.73] * | 13.6 [7.3-34.0] | 0.85 [0.73-0.91] | 3.6 [2.2-7.2] |
| Female | 0.69 [0.59-0.76] | 8.3 [6.9-14.2] | 0.71 [0.56-0.78] * | 9.3 [6.9-20.7] | 0.87 [0.78-0.93] | 3.6 [2.4-6.7] |
| Pathology |  |  |  |  |  |  |
| HS | 0.71 [0.64-0.77] | 7.2[5.4-24.9] | 0.74 [0.71-0.77] | 10.5 [7.4-19.1] | 0.71 [0.86-0.94] | 3.2 [2.0-4.7] |
| non-HS | 0.67 [0.47-0.76] | 9.7 [6.6-16.7] | 0.62 [0.35-0.73] | 12.7 [7.1-36.1] | 0.83 [0.70-0.91] | 3.6 [2.3-7.3] |
| Location of resection |  |  |  |  |  |  |
| Temporal | 0.71 [0.86-0.80] ** | 8.0 [5.7-12.8] ** | 0.70 [0.57-0.77] *** | 10.4 [7.0-22.0] * | 0.87 [0.80-0.93] * | 3.3 [2.2-5.2] |
| Extra-temporal | 0.58 [0.30-0.73] ** | 13.9 [8.2-21.2] ** | 0.52 [0.00-0.71] *** | 23.2 [7.3-68.4] * | 0.80 [0.68-0.90] * | 3.7 [2.8-8.1] |
| Surgical side |  |  |  |  |  |  |
| Left | 0.67 [0.50-0.74] | 9.6 [6.4-16.8] | 0.67 [0.36-0.76] | 13.6 [6.4-43.8] | 0.85 [0.71-0.91] | 3.9 [3.0-7.4] |
| Right | 0.69 [0.53-0.78] | 12.0 [8.0-20.5] | 0.64 [0.41-0.75] | 15.8 [8.3-34.7] | 0.86 [0.77-0.92] | 3.7 [2.4-8.1] |
| MRI field strength |  |  |  |  |  |  |
| 3T | 0.69 [0.51-0.77] | 9.4 [5.9-15.8] | 0.65 [0.37-0.76] | 11.9 [7.0-32.6] | 0.85 [0.74-0.92] | 3.6 [2.2-7.1] |
| 1.5T | 0.56 [0.11-0.64] | 17.0 [8.5-24.4] | 0.58 [0.47-0.69] | 19.3 [11.0-28.3] | 0.82 [0.78-0.89] | 4.0 [3.0-5.4] |
| Isotropy |  |  |  |  |  |  |
| Isotropic | 0.67 [0.50-0.75] | 9.7 [6.1-16.9] | 0.62 [0.36-0.75] * | 12.7 [7.1-32.8] | 0.85 [0.73-0.91] | 3.6 [2.4-7.3] |
| Anisotropic | 0.76 [0.66-0.80] | 10.3 [5.2-18.1] | 0.73 [0.72-0.78] * | 7.8 [6.2-11.6] | 0.67 [0.84-0.93] | 3.2 [1.8-3.9] |
| Mask volume |  |  |  |  |  |  |
| Coefficient (r) | 0.51 *** | 0.01 | 0.35 *** | -0.13 | 0.59 *** | -0.17 |

Performance breakdown of MELD-PostOp across the whole MELD cohort, combining validation predictions and independent test cohorts, is presented in Supplementary Table 4. MELD-PostOp model achieved a consistent median DSC greater than 0.90 across all subgroups except insular resection (DSC=0.81). Median DSCs were comparable between paediatric and adult patients, and between males and females, with no statistically significant differences (corrected *p* > 0.05). Performance varied by pathology, with the highest median DSC observed in HS (DSC=0.98) significantly better than for LEAT, dual pathology, cavernoma, and MCD cases (corrected *p* < 0.001). Median DSC remained high across all resection locations, with the temporal lobe achieving the highest performance (DSC=0.97), followed by the frontal (DSC=0.95) and occipital lobes (DSC=0.93). Temporal lobe resection showed significantly higher DSC than limbic, parietal, and frontal resection (corrected *p* < 0.001). No significant difference was observed between isotropic and anisotropic scans (Supplementary Table 5). A significant positive correlation was observed between the volume of the ground truth resection masks and the DSC (r=0.29, corrected *p* < 0.001) (Supplementary Figure 4), meaning that larger resections had better automated delineation of the cavity. It is worth noting that the DSC is biased against smaller masks, where a single voxel error will have a greater impact. Nevertheless, across the smallest 10% of cavities, the median DSC was still 0.87 (IQR, 0.54-0.94). Furthermore no significant correlation was observed between the volume of ground truth cavities and the HD95 metric. MELD-PostOp achieved a consistent median HD95 lower than 4.0 and showed no significant difference between subgroups, where Epic-CHOP and ResectVol had a significant difference in median HD95 between temporal and extra-temporal resections.

## 4. Discussion

We present MELD-PostOp, an automated, deep learning-based tool designed to enable fast and accurate resection cavity segmentation from postoperative MRI scans in epilepsy patients. MELD-PostOp significantly outperformed two state-of-the-art automated resection cavity masking tools on independent test cohorts, with increased segmentation accuracy, fewer failures and faster evaluation. MELD-PostOp was trained and tested on a large and heterogeneous international multicentre dataset, and performance remained high independent of MRI acquisition parameters, patient age, sex, lesion location, and pathology. MELD-PostOp is a robust, open-source tool that can be used by the field to pursue a range of research questions that aim to understand neuroimaging predictors of seizure freedom and neurocognitive outcomes to ultimately help improve epilepsy surgery outcomes.

### 4.1 Heterogeneous multicentre data for model development and evaluation

MELD-PostOp has been trained and tested on over 1000 postoperative MRI scans from epilepsy centres worldwide. Using a combined independent test set of 137 patients, our DL MELD-PostOp model significantly outperformed current leading methods, which rely on registration and intensity comparisons with the preoperative MRI scan.^8,9^ Methods reliant on differences in intensity between scans exhibit a number of limitations. First, other preoperative to postoperative brain changes, such as changes in ventricle size and postoperative brain shift, may lead to intensity changes and result in inaccurate segmentation of non-resected areas. An example of this is in the segmentation of patient 5’s postoperative resection cavity by Epic-CHOP (Figure 3). Second, these algorithms depend on tissue segmentation differences, and missegmentation of preoperative lesional tissue, for example due to cystic changes being misattributed as cerebrospinal fluid can ultimately lead to undersegmentation of the resection cavity. Third, any misregistration of the postoperative MRI to the preoperative MRI will impact the subsequent segmentation. This is a particular challenge in patients with large resections or young children, where there may be significant brain growth and development between the pre- and postoperative MRI imaging. While these SPM-based intensity models have previously been shown to outperform DL approaches on smaller datasets^16^ the MELD Focal Epilepsies’ large dataset size and diverse resection locations combined with the state-of-the-art nnUNet architecture, likely underpin the significantly improved performance. The inclusion of paediatric cohorts is particularly valuable, as these patients often present with different imaging characteristics, lesion locations and histologies from adults. With the increasing move towards earlier surgery, quantitative postoperative analyses of these cohorts with respect to surgical seizure freedom and neurocognitive outcomes are essential. Moreover, multicentre datasets enable the capture of rarer pathologies and diverse imaging sequences within the training set, thereby enhancing model robustness and generalisability.

### 4.2 Accelerating costly labelling of large datasets through active learning

A major challenge in developing robust segmentation models is the need for large, accurately annotated datasets, which are costly and time-consuming to produce and require expertise. In this study, we adopted an active learning strategy to mitigate this challenge. A prototype model was initially trained on 285 MRI scans with manually annotated masks, and its predictions were quality-controlled and then used to generate labels for an additional 680 scans. This iterative approach substantially reduced the annotation burden while enabling the creation of a large training set. Such strategies demonstrate how active learning can be used in neuroimaging research to accelerate dataset expansion and improve model performance without proportionally increasing expert labelling effort.

### 4.3 Open science

By making MELD-PostOp openly available, we aim to provide the community with a robust tool for postoperative cavity segmentation that can be readily integrated into research workflows. Open-source availability not only facilitates independent replication but also enables benchmarking against existing models, fostering quantitative comparison. Automated and reliable postoperative resection cavity tools combined with large multicentre datasets have the potential to enable big data analysis of neurosurgical outcomes in epilepsy. MELD-PostOp provides a fast, feasible solution for automated masking of large datasets.

### 4.4 Enabling quantitative analysis of post-surgical outcomes

The ability to rapidly, reliably segment large numbers of post-surgical resection cavities could prove invaluable for advancing our understanding of predictors of surgical outcomes. Evidence from recent studies highlights several promising neuroimaging predictors of seizure freedom that need further validation and exploration. For instance, within focal cortical dysplasia type II lesions, ultra-high resolution 7T imaging revealed a “black line sign”, complete resection of which was predictive of postoperative seizure freedom^26^. Other studies have demonstrated that disconnection of thalamocortical white matter tracts in frontal lobe lesions^27^ or diffusion derived grey and white-matter abnormalities in temporal lobe epilepsy^28^ were associated with improved seizure freedom. Resection of the temporal piriform cortex in patients with temporal lobe epilepsy, and particularly in hippocampal sclerosis, has been linked to increased rates of seizure freedom in both adults^29,30^ and children^31^. Beyond seizure outcomes, Spyrantis et al. explored the relationship between the extent of resection and psychobehavioral outcomes in temporal lobe epilepsy. They found that volume of resection was significantly correlated with divided attention, quality of life, and depressive symptoms, whereas sparing of mesial temporal lobe structures for neocortical lesions was associated with fewer depressive symptoms and improved quality of life^4^. Collectively, these studies indicate that our understanding of which lesional features and anatomical structures require resection to reliably achieve seizure freedom remains incomplete. Furthering our understanding could lead to more individualised targeting of procedures that reduce the risk of adverse neurocognitive outcomes. Comprehensive analysis of surgical procedures and post-surgical outcomes in large cohorts will be a key to understanding this complexity, for which accurate and robust automated computational tools such as MELD-PostOp are essential.

### 4.5 Limitations

This study has a number of limitations. First, MELD-PostOp has been trained and tested on only patients who underwent resective surgical procedures, specifically lesionectomies and lobectomies and not interventions such as laser interstitial thermal therapy or disconnection surgeries^32,33^. Independent datasets and models will be required to segment the area of ablated or disconnected tissue. Second, MELD-PostOp has not been evaluated on intraoperative MRI data or MRI data from the initial post-operative period where inflammation, bleeding and swelling may impact its performance. Use on such scans will require careful validation and likely model-retraining on representative data.

### 4.6 Conclusions

MELD-PostOp is an automated tool for segmenting resection cavities from postoperative MRI scans in epilepsy patients. It has improved performance and rapid inference time compared to existing models. The model’s robustness to heterogeneous and unseen data, combined with its minimal input requirements, underscores its suitability for research applications. MELD-PostOp is provided as a fully automated and open-source pipeline to enable large-scale quantitative analysis of postoperative imaging.

## Data Availability

Data sharing of the MELD Focal Epilepsies dataset is restricted as consent was not obtained from participants for public sharing of data.
EPISURG dataset is available at:https://rdr.ucl.ac.uk/articles/dataset/EPISURG_a_dataset_of_postoperative_magnetic_resonance_images_MRI_for_quantitative_analysis_of_resection_neurosurgery_for_refractory_epilepsy/9996158?file=26153588
All code for model design, development and statistical analyses alongside the MELD-PostOp model is available at: https://github.com/MELDProject/MELD-PostOp/tree/main

## Acknowledgements

We would like to thank the undergraduate students Rohan Patel (RP), Beatrice Harrison (BH) and Ellie Nelmes (EN) for their support in creating manual masks of resection cavities, which after quality control and editing, where necessary, were used to train the prototype model.

JS, MR and KW are supported by Wellcome Trust (301991/Z/23/Z) and Epilepsy Research Institute UK (P2208). SA is supported by Epilepsy Research Institute UK(P2208). TJO is supported by NHMRC Investigator Grants (APP1176426 & APP2034258). HX is supported by National Institute of Child Health and Human Development (P50HD105328-01). NTC is supported by the National Institute Of Neurological Disorders And Stroke and the National Institutes of Health (K23NS131522). AGW and AH are supported by Canadian Institute for Health Research, Fonds de recherche du Québec - Santé, Savoy Foundation, Department of surgery at Université de Montréal, CHU Sainte-Justine Foundation. AI is supported by grants from the Fogarty International Center, National Institutes of Health, National Institutes of Aging (R01 AG057234, R01 AG075775, R01 AG21051, R01 AG083799, CARDS-NIH, R01 AG057234), Alzheimer’s Association (SG-20-725707), Rainwater Charitable Foundation, ANID/FONDECYT Regular (1250091 and 1210176 and 1220995), ANID/PIA/ANILLOS ACT210096, JPI JPND-Care, DISCeRN 2025, FONDEF ID20I10152,ANID/FONDAP 15150012; Wellcome Trust (BRAIN-CLIMA 335293/Z/25/Z), and CliCBrain (Horizon ID: 101236426; DOI 10.3030/101236426, Marie Skłodowska-Curie Actions - MSCA). AC is supported by NIHR & GOSH BRC. MHE is supported by The Sigrid Jusélius Foundation. RJP is supported by NIHR & GOSH Children’s Charity. CLY is supported by CNPQ (445340/2024-0/; 313263/2025-6). FC is supported by Sao Paulo Research Foundation (FAPESP) grants 2013/07559-3 and 2021/12956-8. The content of this manuscript is solely the responsibility of the authors and does not necessarily represent the official views of any of the funders. The funders had no role in study design, data collection and analysis, decision to publish or preparation of the manuscript.

This manuscript has been submitted as a preprint to MedRxiv.

