## Supplemental Table / Figure for "Automated Segmentation of Post-Surgical Resection Cavities on MRI in Focal Epilepsy: a MELD Study"

---

#### Supplementary Tables

**Supplementary Table 1.**
Pathological epileptogenic lesion types in the Multicentre Epilepsy Lesion Detection (MELD) Focal Epilepsy (FE) dataset were determined using histolopathological diagnosis. Where histopathological diagnosis was not available, radiological diagnosis was used. For analysis, pathologies were grouped into pathology categories listed below.

| (1) MCD |
|---|
| Focal cortical dysplasia type I (FCD I) |
| Focal cortical dysplasia type IIa (FCD IIa) |
| Focal cortical dysplasia type IIb (FCD IIb) |
| Focal cortical dysplasia type II not otherwise specified (FCD II other) |
| Focal cortical dysplasia type IIIb (FCD IIIb) |
| Focal cortical dysplasia not otherwise specified (FCD other) |
| Malformation of cortical development (MCD) |
| Mild malformation of cortical development (mMCD) |
| Mild malformation of cortical development with oligodendroglial hyperplasia and epilepsy (MOGHE) |
| Polymicrogyria |
| Periventricular nodular heterotopia (PNH) |
| (2) HS |
| Hippocampal sclerosis (HS) / Mesial temporal sclerosis (MTS) |
| Hippocampal gliosis only |
| (3) LEAT |
| Dysembryoplastic neuroepithelial tumour (DNET) |
| Ganglioglioma |
| Astrocytoma |
| Polymorphic low-grade neuroepithelial tumour of the young (PLNTY) |
| Other low-grade tumour |
| Low-grade epilepsy-associated neuroepithelial tumour not otherwise specified (LEAT other) |
| (4) Cavernoma |
| Cavernoma |
| (5) Dual pathology |
| Dual pathology |
| (6) Other pathology |
| Cortical gliosis only |
| Hypothalamic hamartoma (HH) |
| Other pathology |
| Normal |
| (7) Unknown |
| Unknown / Missing data |

**Supplementary Table 2.**
Demographic and clinical characteristics of patients from Stratified Test Cohort (n=50). Median DSC represents the performance of MELD-PostOp.

| | Number of patients (n) | Percentage (%) | Median DSC [IQR] |
|---|---|---|---|
| Age | | | |
| Paediatric | 18 | 36.0 | 0.83 [0.63-0.85] |
| Adults | 30 | 60.0 | 0.86 [0.70-0.89] |
| Unknown | 2 | 4.00 | 0.83 [0.80-0.87] |
| Sex | | | |
| Male | 31 | 62.0 | 0.83 [0.67-0.87] |
| Female | 19 | 38.0 | 0.85 [0.79-0.89] |
| Pathology | | | |
| MCD | 19 | 38.0 | 0.83 [0.68-0.91] |
| HS | 9 | 18.0 | 0.86 [0.83-0.91] |
| LEAT | 9 | 18.0 | 0.77 [0.57-0.88] |
| Cavernoma | 2 | 4.00 | 0.34 [0.21-0.47] |
| Dual pathology | 9 | 18.0 | 0.84 [0.70-0.85] |
| Other | 2 | 4.00 | 0.84 [0.83-0.85] |
| Location of resection | | | |
| Temporal | 32 | 64.0 | 0.85 [0.77-0.89] |
| Frontal | 7 | 14.0 | 0.80 [0.59-0.88] |
| Occipital | 3 | 6.00 | 0.63 [0.60-0.77] |
| Parietal | 4 | 8.00 | 0.83 [0.77-0.87] |
| Limbic | 1 | 2.00 | 0.00 [0.00-0.00] |
| Insular | 1 | 2.00 | 0.71 [0.71-0.71] |
| Other | 2 | 4.00 | 0.89 [0.87-0.91] |
| Surgical side | | | |
| Left | 22 | 44.0 | 0.84 [0.70-0.87] |
| Right | 28 | 56.0 | 0.83 [0.68-0.89] |
| MRI field strength | | | |
| 3T | 42 | 84.0 | 0.84 [0.65-0.89] |
| 1.5T | 8 | 16.0 | 0.82 [0.76-0.88] |
| Isotropy | | | |
| Isotropic | 42 | 84.0 | 0.83 [0.65-0.88] |
| Anisotropic | 8 | 16.0 | 0.86 [0.83-0.94] |

MCD, malformation of cortical development; HS, hippocampal sclerosis; LEAT, low grade epilepsy associated tumour; DSC, dice similarity coefficient; IQR, interquartile range

**Supplementary Table 3.**

Demographic and clinical characteristics of patients from Independent Test Cohort (n=87). Median DSC represents the performance of MELD-PostOp.

| | **Number of patients (n)** | **Percentage (%)** | **Median DSC [IQR]** |
|---|---|---|---|
| Age | | | |
| Paediatric | 15 | 17.2 | 0.89 [0.81-0.94] |
| Adults | 72 | 82.8 | 0.86 [0.75-0.93] |
| Sex | | | |
| Male | 62 | 71.3 | 0.86 [0.75-0.93] |
| Female | 25 | 28.7 | 0.90 [0.78-0.93] |
| Pathology | | | |
| MCD | 37 | 42.5 | 0.87 [0.76-0.93] |
| HS | 12 | 13.8 | 0.92 [0.89-0.94] |
| LEAT | 15 | 17.2 | 0.85 [0.78-0.92] |
| Cavernoma | 8 | 9.20 | 0.75 [0.57-0.90] |
| Dual pathology | 5 | 5.74 | 0.91 [0.91-0.94] |
| Other | 8 | 9.20 | 0.74 [0.55-0.90] |
| Unknown | 2 | 2.90 | 0.76 [0.69-0.84] |
| Location of resection | | | |
| Temporal | 51 | 58.6 | 0.91 [0.81-0.94] |
| Frontal | 22 | 25.3 | 0.81 [0.76-0.90] |
| Occipital | 2 | 2.30 | 0.90 [0.90-0.91] |
| Parietal | 10 | 11.5 | 0.78 [0.68-0.86] |
| Limbic | 1 | 1.15 | 0.63 [0.63-0.63] |
| Other | 1 | 1.15 | 0.61 [0.61-0.61] |
| Surgical side | | | |
| Left | 43 | 49.4 | 0.85 [0.71-0.93] |
| Right | 44 | 50.6 | 0.89 [0.78-0.93] |
| MRI field strength | | | |
| 3T | 85 | 97.7 | 0.87 [0.76-0.93] |
| 1.5T | 2 | 2.30 | 0.86 [0.84-0.88] |
| Isotropy | | | |
| Isotropic | 85 | 97.7 | 0.87 [0.76-0.93] |
| Anisotropic | 2 | 2.30 | 0.85 [0.81-0.88] |

MCD, malformation of cortical development; HS, hippocampal sclerosis; LEAT, low grade epilepsy associated tumour; DSC, dice similarity coefficient; IQR, interquartile range

**Supplementary Table 4.**
MELD-PostOp model subgroup performance across clinical and imaging variables evaluated on the entire MELD cohort (n=969).

| | **Median DSC [IQR]** |
|---|---|
| Age | |
| Paediatric | 0.95 [0.87-0.98] |
| Adult | 0.96 [0.88-0.98] |
| Sex | |
| Male | 0.94 [0.86-0.98] |
| Female | 0.96 [0.90-0.98] |
| Pathology | |
| MCD | 0.94 [0.86-0.98] |
| HS | 0.98 [0.95-0.99] |
| LEAT | 0.94 [0.87-0.98] |
| Cavernoma | 0.92 [0.66-0.98] |
| Dual pathology | 0.96 [0.88-0.98] |
| Other | 0.92 [0.82-0.95] |
| Unknown | 0.96 [0.92-0.97] |
| Location of resection | |
| Temporal | 0.97 [0.90-0.98] |
| Frontal | 0.95 [0.85-0.98] |
| Occipital | 0.93 [0.85-0.97] |
| Parietal | 0.93 [0.79-0.97] |
| Limbic | 0.90 [0.68-0.95] |
| Insular | 0.81 [0.61-0.93] |
| Other | 0.93 [0.85-0.97] |
| Surgical side | |
| Left | 0.95 [0.87-0.98] |
| Right | 0.96 [0.88-0.98] |
| MRI field strength | |
| 3T | 0.96 [0.88-0.98] |
| 1.5T | 0.94 [0.86-0.97] |
| Isotropy | |
| Isotropic | 0.95 [0.86-0.98] |
| Anisotropic | 0.97 [0.92-0.98] |

MCD, malformation of cortical development; HS, hippocampal sclerosis; LEAT, low grade epilepsy associated tumour; DSC, dice similarity coefficient; IQR, interquartile range

**Supplementary Table 5.**

Subgroup comparisons of MELD-PostOp performed across variables (n=969). Multiple comparison correction was applied using Bonferroni adjustment for subgroup level analyses (n=8). For categorical variables with significant overall group differences, pairwise comparisons were conducted with separate p value correction applied within each variable (pathology, 21 pairwise comparisons; location of resection, 21 pairwise comparisons).

| | **Comparison** | ***p*** | **Corrected *p*** | **Sig.** | **Sig. corr.** |
|---|---|---|---|---|---|
| 1 | Age | 9.981E-01 | 1.000E+00 | ns | ns |
| 2 | Sex | 9.989E-01 | 1.000E+00 | ns | ns |
| 3 | Pathology | 3.732E-16 | 2.986E-15 | *** | *** |
| 3-1 | MCD-HS | 5.506E-16 | 1.156E-14 | *** | *** |
| 3-2 | MCD-LEAT | 5.890E-01 | 1.000E+00 | ns | ns |
| 3-3 | MCD-Cavernoma | 5.053E-02 | 1.000E+00 | ns | ns |
| 3-4 | MCD-Dual pathology | 1.803E-01 | 1.000E+00 | ns | ns |
| 3-5 | MCD-Other | 8.304E-02 | 1.000E+00 | ns | ns |
| 3-6 | MCD-Unknown | 6.849E-01 | 1.000E+00 | ns | ns |
| 3-7 | HS-LEAT | 2.916E-11 | 6.124E-10 | *** | *** |
| 3-8 | HS-Dual pathology | 2.282E-05 | 4.792E-04 | *** | *** |
| 3-9 | HS-Cavernoma | 5.398E-08 | 1.134E-06 | *** | *** |
| 3-10 | HS-Other | 2.532E-06 | 5.317E-05 | *** | *** |
| 3-11 | HS-Unknown | 4.272E-02 | 8.971E-01 | * | ns |
| 3-12 | LEAT-Dual pathology | 3.528E-01 | 1.000E+00 | ns | ns |
| 3-13 | LEAT-Cavernoma | 4.052E-02 | 8.509E-01 | * | ns |
| 3-14 | LEAT-Other | 5.784E-02 | 1.000E+00 | ns | ns |
| 3-15 | LEAT-Unknown | 7.605E-01 | 1.000E+00 | ns | ns |
| 3-16 | Cavernoma-Dual pathology | 1.828E-02 | 3.839E-01 | * | ns |
| 3-17 | Cavernoma-Other | 9.956E-01 | 1.000E+00 | ns | ns |
| 3-18 | Cavernoma-Unknown | 3.815E-01 | 1.000E+00 | ns | ns |
| 3-19 | Dual pathology-Other | 2.190E-02 | 4.599E-01 | * | ns |
| 3-20 | Dual pathology-Unknown | 8.476E-01 | 1.000E+00 | ns | ns |
| 3-21 | Other-Unknown | 1.660E-01 | 1.000E+00 | ns | ns |
| 4 | Location of resection | 2.311E-11 | 1.849E-10 | *** | *** |
| 4-1 | Frontal-Temporal | 6.980E-06 | 1.466E-04 | *** | *** |
| 4-2 | Frontal-Parietal | 6.649E-02 | 1.000E+00 | ns | ns |
| 4-3 | Frontal-Occipital | 7.897E-01 | 1.000E+00 | ns | ns |
| 4-4 | Frontal-Limbic | 2.575E-03 | 5.408E-02 | ** | ns |
| 4-5 | Frontal-Insular | 3.253E-02 | 6.831E-01 | * | ns |
| 4-6 | Frontal-Other | 4.248E-01 | 1.000E+00 | ns | ns |
| 4-7 | Temporal-Parietal | 9.831E-07 | 2.065E-05 | *** | *** |
| 4-8 | Temporal-Occipital | 3.790E-02 | 7.959E-01 | * | ns |
| 4-9 | Temporal-Limbic | 3.690E-06 | 7.749E-05 | *** | *** |
| 4-10 | Temporal-Insular | 4.879E-03 | 1.025E-01 | ** | ns |
| 4-11 | Temporal-Other | 1.687E-02 | 3.543E-01 | * | ns |
| 4-12 | Parietal-Occipital | 3.922E-01 | 1.000E+00 | ns | ns |
| 4-13 | Parietal-Limbic | 8.281E-02 | 1.000E+00 | ns | ns |
| 4-14 | Parietal-Insular | 1.491E-01 | 1.000E+00 | ns | ns |
| 4-15 | Parietal-Other | 8.911E-01 | 1.000E+00 | ns | ns |
| 4-16 | Occipital-Limbic | 5.203E-02 | 1.000E+00 | ns | ns |
| 4-17 | Occipital-Insular | 8.997E-02 | 1.000E+00 | ns | ns |
| 4-18 | Occipital-Other | 6.992E-01 | 1.000E+00 | ns | ns |
| 4-19 | Limbic-Insular | 6.053E-01 | 1.000E+00 | ns | ns |
| 4-20 | Limbic-Other | 9.973E-02 | 1.000E+00 | ns | ns |
| 4-21 | Insular-Other | 2.045E-01 | 1.000E+00 | ns | ns |
| 5 | Surgical side | 9.272E-01 | 1.000E+00 | ns | ns |
| 6 | MRI field strength | 4.228E-03 | 3.383E-02 | ** | * |
| 7 | Isotropy | 9.978E-01 | 1.000E+00 | ns | ns |
| 8 | Resection volume | 1.050E-19 | 8.398E-19 | *** | *** |

Sig., significance; Sig. corr., significance after bonferroni adjustment; LH, left hemisphere; RH, right hemisphere; MCD, malformation of cortical development; HS, hippocampal sclerosis; LEAT, low grade epilepsy associated tumour; *, p <0.05; **, P< 0.01; ***, p<0.001; ns, not significant

### Supplementary Figures

**Supplementary Figure 1.**
Representative 3*14 visual quality control (QC) image from a case that passed QC.

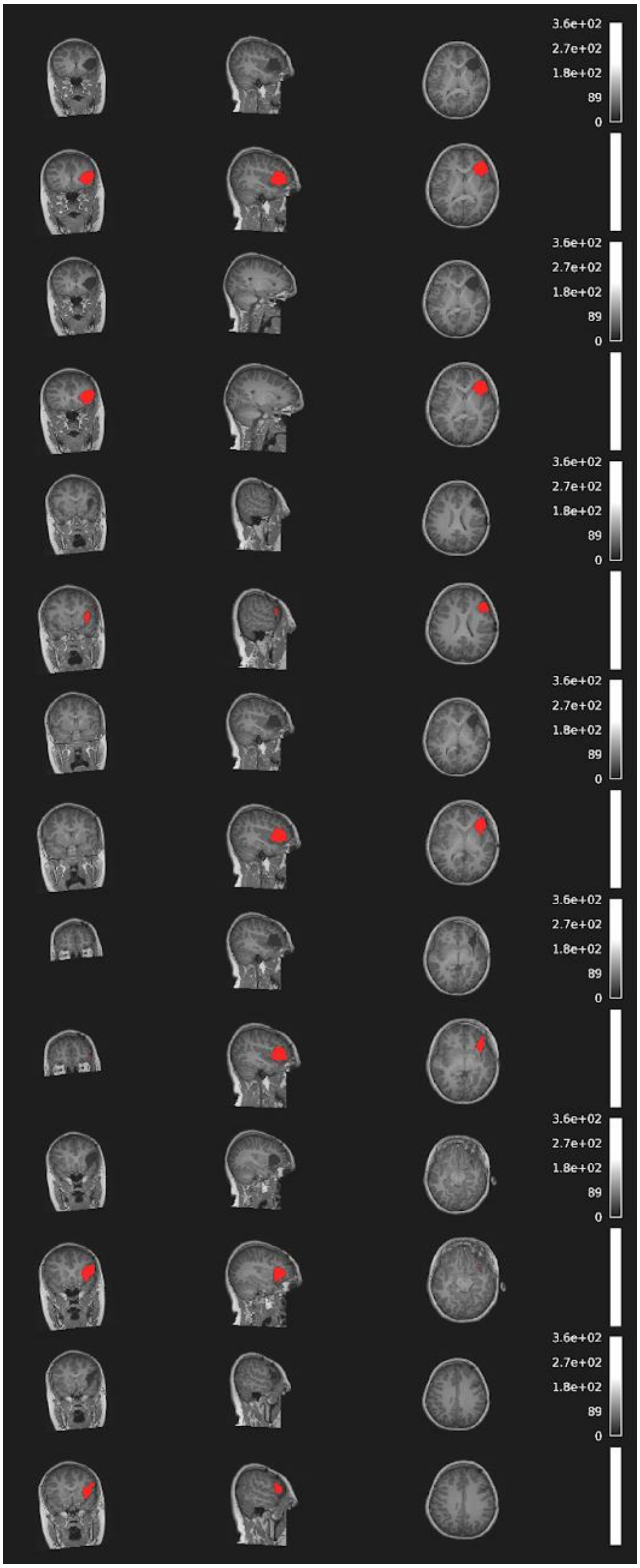

**Supplementary Figure 2.**
Representative 3*14 visual quality control (QC) image from a case that failed QC. The resection cavity segmentation was incomplete and subsequently corrected through manual editing.

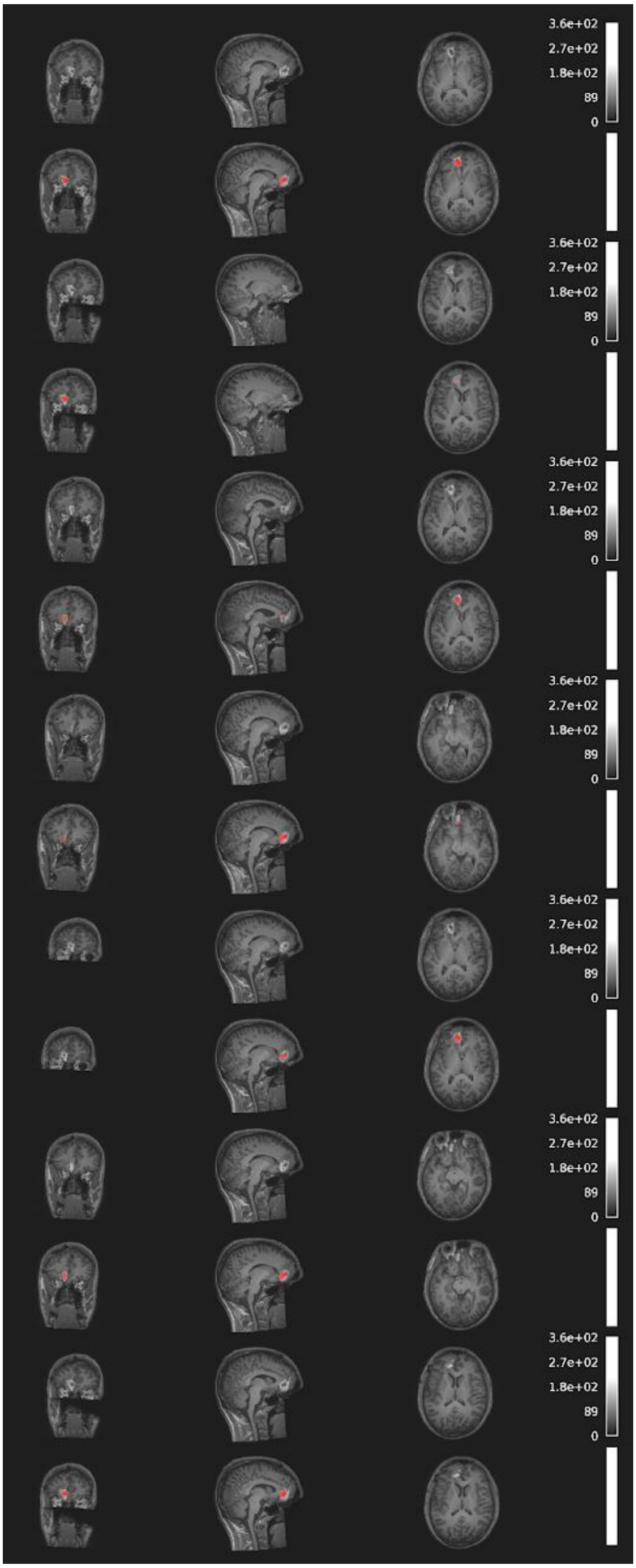

**Supplementary Figure 3.**

Raincloud plots illustrate change in dice similarity coefficient (DSC) for cases that failed QC after initial inference with the Prototype model (n=85), comparing Prototype and MELD-PostOp performance. Paired values depict within-case changes in segmentation accuracy. Overall DSC increased with MELD-PostOp, indicating improved segmentation performance. Median DSC increased from 0.31 (IQR: 0.00-0.99) for the Prototype model to 0.70 (IQR: 0.28-0.93) for MELD-PostOp. However, this difference was not statistically significant (Mann-Whitney U test, p =0.143).

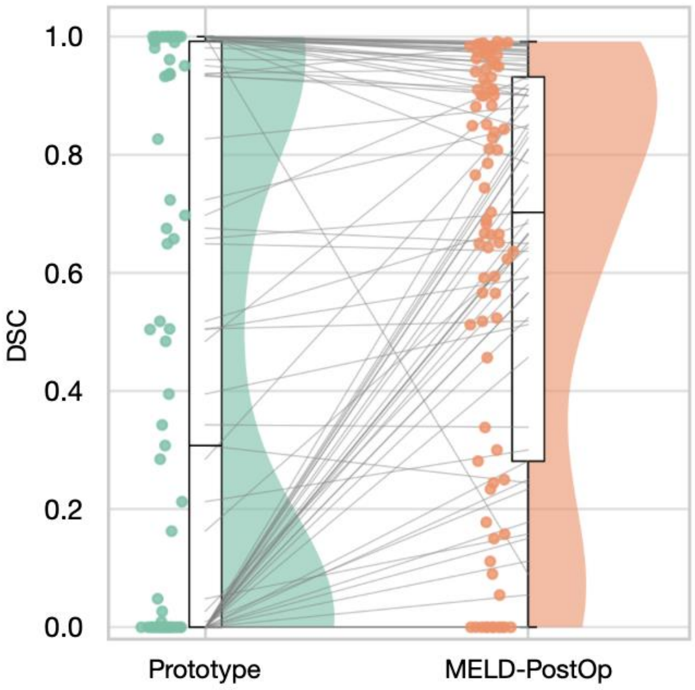


**Supplementary Figure 4.**

Spearman's rank correlation between manual resection cavity volume and dice similarity coefficient (DSC) for manual versus MELD-PostOp predictions (n=969). A statistically significant positive correlation was observed (r=0.286, $p < 0.001$), indicating higher DSC with increasing cavity volume. The mean manual mask volume was 17.45 $cm^3$, with a median of 13.39 $cm^3$.

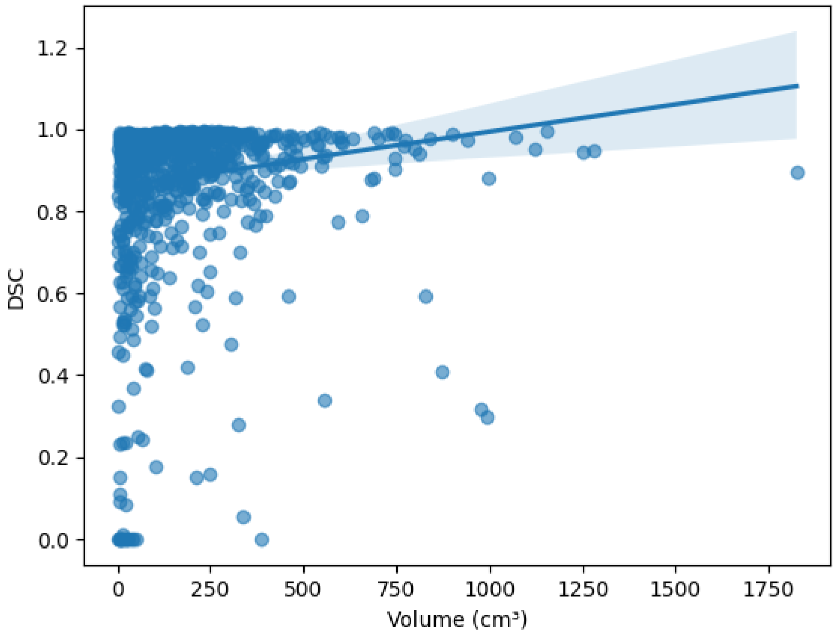

**Supplementary Figure 6.**
Runtime analysis of Epic-CHOP, ResectVol, and MELD-PostOp were performed on the same 10 subjects using identical computational hardware. Epic-CHOP demonstrated a median runtime of 3363.5s, ResectVol 659.0s, and MELD-PostOp 15.0s. Significant differences were observed between all three models ($p < 0.001$). Mean runtime was 3205s, 612s, and 17s, respectively.

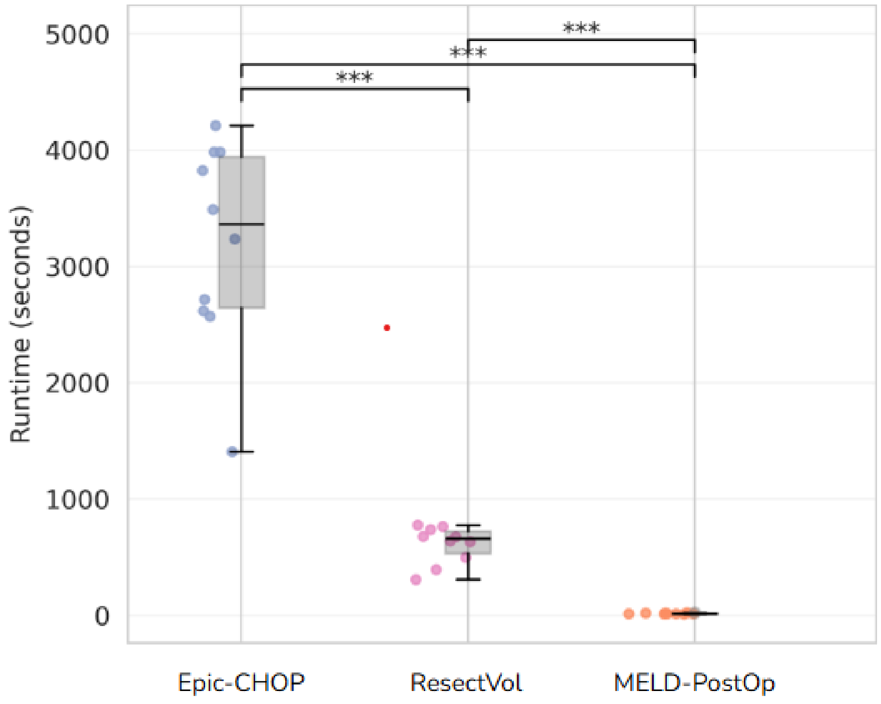
